# Objective turning measures improve diagnostic accuracy and relate to real-world mobility/combat readiness in chronic mild traumatic brain injury

**DOI:** 10.1101/2024.03.11.24304109

**Authors:** Peter C. Fino, Prokopios Antonellis, Lucy Parrington, Margaret M. Weightman, Leland E. Dibble, Mark E. Lester, Carrie W. Hoppes, Laurie A. King

**Affiliations:** Department of Health & Kinesiology, University of Utah, Salt Lake City, Utah, United States of America; Department of Neurology, Oregon Health & Science University, Portland, Oregon, United States of America; Department of Dietetics, Human Nutrition and Sport, La Trobe University, Melbourne, Victoria, Australia; Courage Kenny Research Center-Allina Health, Minneapolis, Minnesota, United States of America; Department of Physical Therapy & Athletic Training, University of Utah, Salt Lake City, Utah, United States of America; Department of Physical Therapy, University of Texas Rio Grande Valley, Edinburg, Texas, United States of America; Army-Baylor University Doctoral Program in Physical Therapy, Fort Sam Houston, Texas, United States of America; National Center for Rehabilitative Auditory Research, Veterans Affairs Portland Health Care System, Portland, Oregon, United States of America

**Keywords:** concussion, mild traumatic brain injury, assessment, turning measures

## Abstract

**Introduction:** Balance and mobility problems are common consequences after mild traumatic brain injury (mTBI). However, turning and non-straight locomotion, which are required for daily living, are rarely assessed in clinical tests of function after mTBI. Therefore, the primary goals of this study were to assess 1) the added value of clinic-based turning task variables, obtained using wearable sensors, over standard general assessments of mobility, and 2) assess the associations between general assessments of mobility, objective variables from clinic-based turning tasks, and ecologically-relevant functional tasks.

**Materials and Methods:** Fifty-three individuals with mTBI and 57 healthy controls participated across three sites. Participants were tested in a single session that encompassed self-reported questionnaires including demographic information and balance and mobility testing including the use of wearable sensors. Lasso regression models and the area under the receiver-operator characteristic curve (AUC) assessed diagnostic accuracy. Partial correlation coefficients assessed the relationship between each variable with ecologically-relevant functional tasks.

**Results:** Multivariate models revealed high diagnostic accuracy, with an AUC of 0.92, using multiple clinic-based turning variables. The complex turning course (CTC) yielded the highest multivariate AUC (95% CI) of 0.90 (0.84, 0.95) for single task, and the average lap time from the CTC had the highest univariate AUC (95% CI) of 0.70 (0.58, 0.78). Turning variables provided added value, indicated by higher AUCs, over standard general assessments of mobility. Turning variables had strong associations with ecologically-relevant functional tasks and outperformed general assessments of mobility.

**Discussion:** Clinic-based turning tasks, especially the CTC, have high diagnostic accuracy, strong associations with ecologically-relevant functional tasks, and require relatively short time(s) to complete. Compared to general assessments of mobility, clinic-based turning tasks may be more ecologically-relevant to daily function. Future work should continue to examine the CTC alongside other promising tools for return-to-activity assessments.

## INTRODUCTION

Balance and mobility problems are common consequences after mild traumatic brain injury (mTBI), with variable presentations depending on the specific cognitive, motor, and sensory demands of the task as well as personal and injury factors of the individual with mTBI (1, 2). Consequently, results from objective assessments of balance and mobility are important clinical tools that can inform rehabilitation prescription and track recovery over time. Capturing deficits after mTBI and understanding their impact on a patient’s life requires assessments of mobility that reflect the demands of daily living. While extensive literature on dual-task (DT) gait, where mobility tasks are combined with a simultaneous cognitive task to mirror everyday life, demonstrates diagnostic utility in people with mTBI (3–8), these tasks are often limited to straight-line walking and artificial, laboratory-based cognitive tasks such as serial 3 subtraction. Other multi-faceted clinical assessment batteries such as the Functional Gait Assessment (FGA) (9), High-level Mobility Assessment Tool (HiMAT) (10, 11), 4-Item Hybrid Assessment of Mobility for mTBI (HAM-4-mTBI) (12), and mini Balance Evaluation Systems Test (mini-BESTest) (13) include a variety of mobility tasks, such as walking with horizontal head turns, running, bounding, and reactive stepping with varying motor demands. The majority of individual test items within these general measures of mobility similarly focus on straight-line walking. Non-straight locomotion and ambulatory turning to navigate complex environments are rarely assessed in clinical tests of function after mTBI. Recognizing that work has begun to identify more complex and ecologically relevant tasks such as the Assessment of Military Multitasking Performance (AMMP) (14, 15), and more recently the Portable Warrior Test of Tactical Agility (POWAR-TOTAL) (16) that may contribute to return to duty decisions in the military, tasks that identify issues with key components of everyday mobility such as those involving turning may provide important discriminatory ability for individuals with persistent deficits affecting function in a targeted fashion.

Ambulatory turning is an important characteristic of daily mobility as individuals inevitably must navigate through complex environments that do not permit straight-line travel. Approximately 40% of all steps are non-straight steps involving some degree of turning (17). Stable turning requires anticipatory postural control (18), asymmetrical loading across limbs (19), and dynamic reweighting of sensory information to account for time-varying gravitoinertial accelerations (20, 21). As turning is often enacted to get to a target object or location, people also reorient their gaze and stabilize visual information using sophisticated oculomotor and vestibulo-ocular reflexes, and use sequential rotations of the head, trunk, pelvis, and feet to reorient to the new direction of travel (22–24). These characteristics of turning are unique from straight gait, and models of mobility should include turning as a factor that is independent from other traditional measures of gait such as pace, rhythm, and variability (25, 26).

Preliminary work in a sample of individuals with persisting balance-related symptoms after mTBI (i.e., chronic mTBI) demonstrated slower turning speeds and more variable head-on-body coordination when walking along a complex turning course simulating turns performed in daily life (27). Other studies have reported abnormal balance control during turning in otherwise asymptomatic athletes recovering from mTBI (28). When selecting an optimized set of clinical items from the FGA and HiMAT, the Gait with Pivot Turn test item from the FGA was one of only four test items retained for use in populations with chronic mTBI (12). These studies suggest assessments of turning may have clinical value in populations with mTBI. However, such results leave ambiguity over which assessment of turning, and which variables, are most relevant for assessing and monitoring people with mTBI. The clinical value of turning may depend on the specific demands, instructions, and outcomes of the turning task. For example, tasks requiring faster turning speeds can elicit more severe symptoms due to greater rates of change in visual and vestibular sensory stimulation (29–31), and this provocation of symptoms may affect performance. Other methodological considerations, like the sharpness of the turn angle (27), the height of objects such as cones versus lines on the ground outlining the course (32), or the cognitive complexity and modality of the task (33), can similarly affect turning behavior, such as turning speed and head-body coordination. These factors may affect the clinical value of turning measures for people with mTBI.

The need for objective measurements of turning becomes evident due to the limitations of self-report questionnaires and the often subtle and diverse ways mTBI can manifest (4, 34–38). For instance, one of the most common scales used to assess balance following mTBI is the Balance Error Scoring System (BESS), in which healthcare and sports medicine professionals subjectively count errors and instances of loss of balance while the patient assumes various stance positions with their eyes closed (39). However, even when the BESS results appear normal, more objective measures such as the instrumented BESS (utilizing a single inertial sensor during the testing protocol) may reveal abnormalities (35, 36). Similarly, instrumented sway from the Clinical Test for Sensory Interaction in Balance (mCTSIB) reveal abnormalities where clinical ratings show normal function after mTBI (40). Clinical scales that include turning, such as the Berg Balance Scale (41), often have ceiling effects and may not detect subtle deficits after mTBI especially in highly fit and athletic populations (34). In contrast, instrumented measures of turning, such as peak velocity and segmental coordination across various turn angles, provide reliable measures capable of detecting subtle deficits without relying on subjective visual ratings (42). Both the Veterans Affairs/Department of Defense (VA/DoD) Clinical Practice Guideline for the Management and Rehabilitation of Post-acute Mild Traumatic Brain Injury(43) and Sixth Consensus Statement on Concussion in Sport (44) recommend multimodal screenings that include gait, with the VA/DoD guidelines specifically calling for the evaluation of “walking, tandem walking, walking with head turns, and whole-body turning.” Official evaluation of sport-related concussion should include “timed tandem gait as a single task and a more complex dual-task with the addition of a cognitive task (such as serial 7’s, months backwards or word recall backwards)” (44). The clinical recommendations for including gait, especially turning and DT measures, in mTBI screenings underscores the importance of such measurements, and emphasizes the need for objective tools that are able to detect residual and subtle deficits.

Beyond diagnostic accuracy, the degree to which objective measures of turning reflect a patient’s ability to return-to-duty (RTD), work, or sport remains unclear. This association with real-world function is particularly relevant for rehabilitation decisions that must determine whether subtle residual deficits after mTBI impact one’s readiness for duty, work, or sport. In military populations, where an individual must be able to perform warrior tasks and battle drills such as moving under fire, reacting to contact, and maintaining situational awareness, performance on such duty-relevant tasks is essential to maintain combat effectiveness and ensure survival for themselves and their fellow service members. Common general measures of mobility such as the FGA and HiMAT may not represent the demands imposed by daily life, sports, or specific warrior tasks (urban assault, movement to contact, etc.) that require complex multi-segmental coordination while under cognitive load (45, 46). The clinical utility of turning metrics is jointly determined by the diagnostic accuracy and association with real-world mobility and combat readiness.

Therefore, the first goal of this larger study (37) was to assess diagnostic accuracy—the added value of objective turning measures over standard, general assessments of mobility in identifying concussed individuals who are not fully recovered. We expected that variables from clinic-based turning tasks would have better diagnostic accuracy for identifying individuals with mTBI, as indicated by a greater area under the receiver-operator characteristic (ROC) curve (AUC) compared to general measures of mobility. The second goal of this study was to support the validity of the turning variables and assess the associations between general measures of mobility and clinic-based turning tasks with performance in a community ambulatory task (CAT) and a military-relevant simulated urban patrol task (SUP) which we are designating as ecologically-relevant functional tasks. We hypothesized that objective turning variables obtained during the clinic-based turning tasks would be more strongly associated with performance in ecologically-relevant functional tasks compared to performance in general measures of mobility. Finally, we sought to provide recommendations on the best turning tasks and variables for future clinical implementation based on the diagnostic accuracy, added value, and association with the ecologically-relevant functional tasks.

## MATERIALS and METHODS

### Participants

As part of the ReTURN study protocol (ClinicalTrials.gov: NCT03892291) (37), a total of 53 individuals with mTBI and 57 healthy controls (Table 1) were recruited from May 15, 2019 to October 20, 2020 across three sites (Oregon Health & Science University, Portland, OR; University of Utah, Salt Lake City, UT; Courage Kenny Research Center - Allina Health, Minneapolis, MN). Inclusion criteria for those with mTBI were: (1) have a diagnosis of mTBI, (2) be between 18 and 50 years of age, and (3) be outside of the acute stage (>3 weeks post-concussion) but within 3 years of their most recent mTBI and still reporting symptoms. Control participants either had no history of mTBI or were more than 7 years removed from their most recent mTBI and had no reported residual symptoms. Potential participants were excluded if they: (1) had a history of any other injury, medical condition, or neurological illness that could potentially impair their balance (i.e., lower extremity injury, recent surgery, stroke), (2) had a current substance abuse disorder, (3) were pregnant, or (4) were unable to communicate in English. The study was conducted in accordance with the Declaration of Helsinki (1964) and approved by the Institutional Review Board at each of the sites. Informed written consent was obtained prior to participation.

**Table 1.**
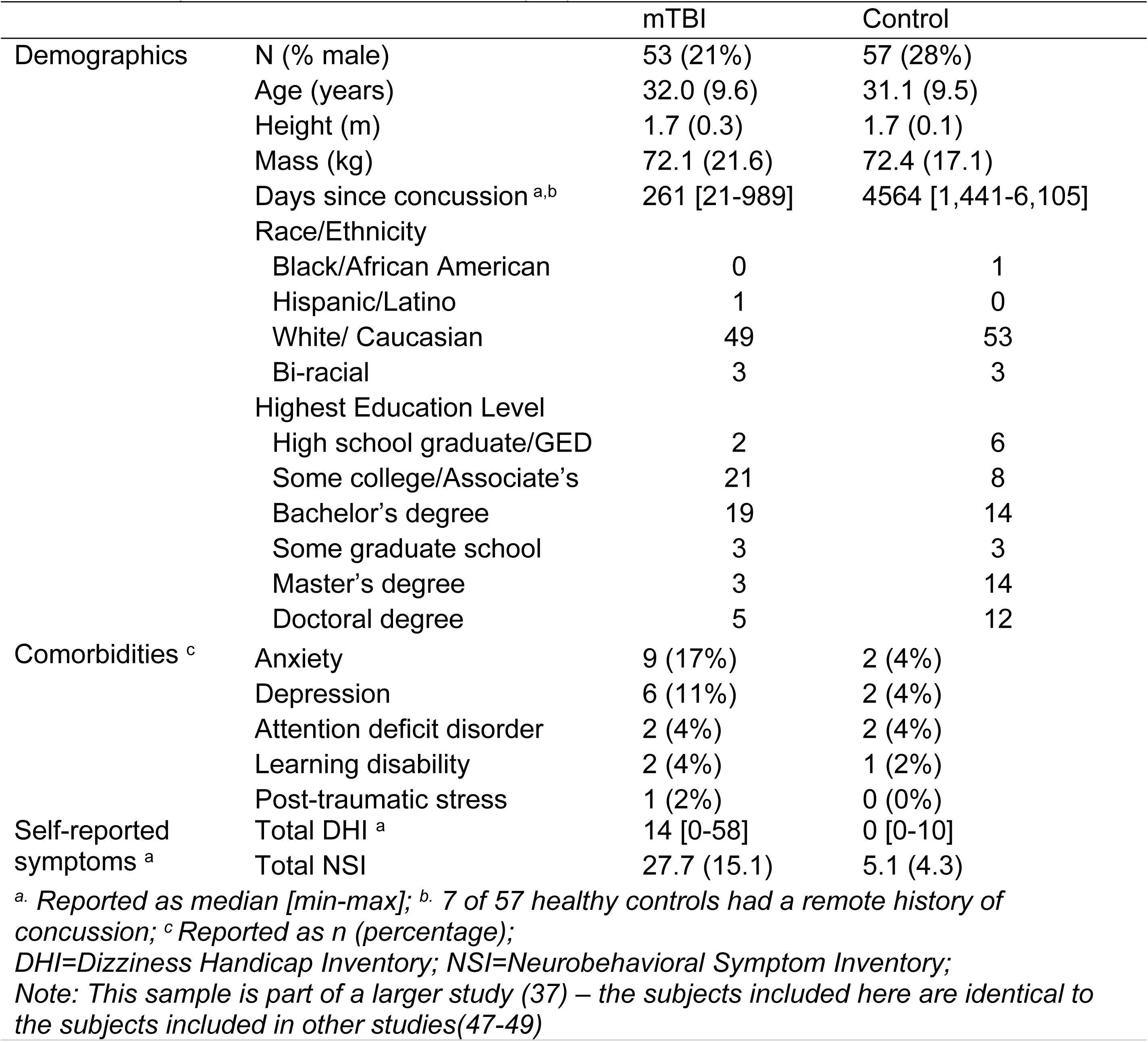
Demographic characteristics for 53 individuals with mild traumatic brain injury (mTBI) and 57 healthy controls reported as mean (SD) unless otherwise noted.

|  |  | mTBI | Control |
| --- | --- | --- | --- |
| Demographics | N (% male) | 53 (21%) | 57 (28%) |
|  | Age (years) | 32.0 (9.6) | 31.1 (9.5) |
|  | Height (m) | 1.7 (0.3) | 1.7 (0.1) |
|  | Mass (kg) | 72.1 (21.6) | 72.4 (17.1) |
|  | Days since concussion <sup>a,b</sup> | 261 [21-989] | 4564 [1,441-6,105] |
|  | Race/Ethnicity |  |  |
|  | Black/African American | 0 | 1 |
|  | Hispanic/Latino | 1 | 0 |
|  | White/ Caucasian | 49 | 53 |
|  | Bi-racial | 3 | 3 |
|  | Highest Education Level |  |  |
|  | High school graduate/GED | 2 | 6 |
|  | Some college/Associate's | 21 | 8 |
|  | Bachelor's degree | 19 | 14 |
|  | Some graduate school | 3 | 3 |
|  | Master's degree | 3 | 14 |
|  | Doctoral degree | 5 | 12 |
| Comorbidities <sup>c</sup> | Anxiety | 9 (17%) | 2 (4%) |
|  | Depression | 6 (11%) | 2 (4%) |
|  | Attention deficit disorder | 2 (4%) | 2 (4%) |
|  | Learning disability | 2 (4%) | 1 (2%) |
|  | Post-traumatic stress | 1 (2%) | 0 (0%) |
| Self-reported symptoms <sup>a</sup> | Total DHI <sup>a</sup> | 14 [0-58] | 0 [0-10] |
|  | Total NSI | 27.7 (15.1) | 5.1 (4.3) |

<sup>a</sup>. Reported as median [min-max]; <sup>b</sup>. 7 of 57 healthy controls had a remote history of concussion; <sup>c</sup> Reported as n (percentage); DHI=Dizziness Handicap Inventory; NSI=Neurobehavioral Symptom Inventory; Note: This sample is part of a larger study (37) – the subjects included here are identical to the subjects included in other studies(47-49)

### Procedures

Participants completed one testing session that encompassed self-reported questionnaires including demographic information and symptom checklists, neurocognitive testing, and balance and mobility testing including wearable sensors. For the primary purposes of this study, only the mobility procedures are further described in detail.

#### Clinic-Based Turning Tasks

Participants completed three clinic-based turning tasks in a randomized order (Figure 1): 1) a one-minute walk test (1MW) that involved walking at a comfortable pace between two lines on the ground marked 6 m apart and included 180° turns, 2) a modified Illinois Agility Test (mIAT) that involved running at a maximal safe speed around cones with multiple turns (end, slalom, and mid), and 3) a 140-second walk around a complex turning course (CTC) designed to mimic turns performed in daily life, which involved walking at a comfortable pace around 45°, 90°, and 135° turns. Each of the three turning tasks were completed twice (once under single-task (ST) and another time under DT conditions). The cognitive component for the 1MW and mIAT DT conditions was an 8-digit alpha-numeric grid coordinate memorization task that was introduced within the context of a civilian geocaching activity (15). The cognitive overlay for the CTC DT condition involved monitoring and responding to keywords in a custom-developed simulated radio chatter task designed to mimic demands of military service (14, 15).

**Fig 1.**
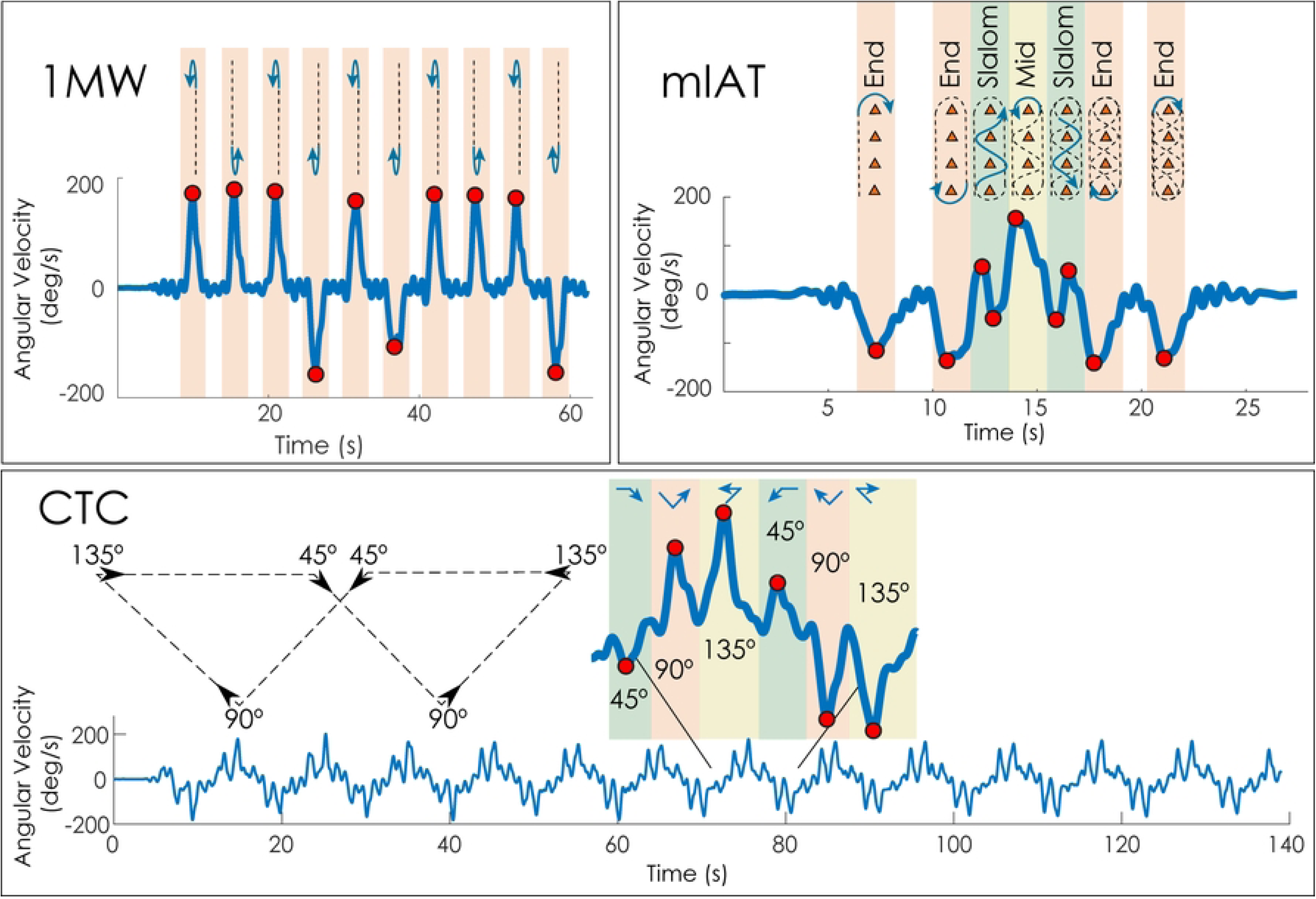
Schematic of the three clinic-based turning tasks with individual turns marked. Turn angles for the complex turning course (CTC) are with respect to straight walking.

#### General Measures of Mobility

In addition to the clinic-based turning tasks, participants completed two standard clinical assessments to obtain general measures of mobility including the FGA (9) and the HiMAT (10). These two clinical assessments were completed and scored based on standard instructions.

#### Ecologically-Relevant Functional Tasks

The Community Ambulatory Task (CAT) involved participants walking at their self-selected pace while navigating through a building following landmark-based directions (e.g., “walk down the hallway towards the black doors”). Instructions were provided verbally by a researcher who walked behind the participant to avoid affecting the pace of the participant. The CAT was unique to each site, but all took approximately 6-7 minutes to complete and contained a standardized set of components common to everyday ambulation (i.e., turns, stairs, use of signage).

The Simulated Urban Patrol (SUP) involved navigating a small subdivided room containing LED targets representing hostile (red) and friendly (blue) targets. The room was constructed using PVC pipe and 2.44 m tall black curtains. The LED targets were constructed using an Adafruit Circuit Playground Express circuit board (Adafruit Industries, LLC; New York, NY) programmed to display red or blue LEDs and to respond to an infrared LED signal from a laser-tag weapon (Model T1504, Dynasty Toys). Upon ‘tagging’ the targets with the laser-tag weapon, the targets were programmed to turn off. If a friendly target was tagged, the target was programmed to turn white. Three hostile targets were programmed to require multiple shots to fully clear – the target would illuminate 2/3 of the red LEDs (1/3 cleared) after one shot, 1/3 of the red LEDs (2/3 cleared) after two shots, and no LEDs (fully cleared) after three shots. Ten total targets were displayed in the subdivided room.

Using a laser-tag weapon, participants were instructed to clear all hostile targets as quickly as possible without tagging friendly targets. Participants were given one practice trial where all targets were set to hostile (red). Following the practice trial, three SUP trials were completed and recorded. The location of the targets was fixed throughout the test, but the configuration of each target (hostile vs. friendly) was changed between trials.

### Data Analysis and Outcome Measures

#### Clinic-Based Turning Tasks

During all clinic-based turning tasks, inertial measurement units (128 Hz; APDM, a Clario Company, Portland, OR, USA) on the forehead, sternum, lumbar spine, and bilateral feet collected tri-axial acceleration and angular velocity data. During each of the turning tasks, peak angular rates for each segment, segmental coordination, and overall speed (e.g., lap times) were obtained from the inertial sensors using previously defined algorithms (42, 50). Briefly, body-fixed yaw angular velocities were extracted from each 1MW, mIAT, and CTC test and filtered using a 1.5 Hz low-pass phaseless Butterworth filter. Specific turns for the mIAT and CTC were identified using a template-based approach based on the prescribed path (50), and each turn variable was matched to each turn type (slalom, mid, and end turns for mIAT; 45°, 90°, and 135° turns for CTC). Speed variables (PeakHeadV, PeakTrunkV, PeakLumbarV) were defined as the peak angular rate of each segment (Head, Trunk, Pelvis, respectively). Intersegmental coordination variables (Lumbar2Head, Lumbar2Trunk, Trunk2Head) were defined as the difference in time between peak angular rates of two segments where positive values indicate the superior segment led the inferior segment (e.g., a Lumbar2Head value of +100 indicates the head reached its peak velocity 100 ms before the pelvis reached its peak velocity for a given turn type) (27). Since each task included multiple turns across each turn angle, variables were averaged within each turn angle. In addition to these measures, the task completion time for the mIAT, the average lap time for the CTC, and the variability (standard deviation) of lap time for the CTC were retained as variables. Therefore, all processing yielded a total of 6 variables (3 speed + 3 coordination) for the 1MW, 19 variables (9 speed + 9 coordination + 1 completion time) for the mIAT, and 20 variables (9 speed + 9 coordination + 2 lap times) for the CTC. The greater number of variables for the mIAT and CTC was due to three different turn angles compared to only one turn angle (180°) for the 1MW.

Additional exploratory variables were also examined for each task. These exploratory variables included measures of head turn symmetry, range of motion, and variabilities of peak turning speed and intersegmental coordination. Head turn symmetry was defined as the ratio of peak turning speed of the head during turns to the left divided by peak turning speed of the head during turns to the right. Head range of motion was defined as the difference between the 95^th^ percentile and 5^th^ percentile of the head-on-trunk angle, obtained through cumulative trapezoidal integration (c*umtrapz* function in MATLAB) of the head-on-trunk angular velocity, over the duration of the entire trial. The variabilities of peak turning speed and intersegmental coordination variables were defined as the standard deviation of each outcome within a given trial. This yielded a total of 152 different turning variables. A list and description of all variables is provided in the supplemental material (Supplement A).

#### General Measures of Mobility

Clinical outcomes of FGA total score and HiMAT total score were calculated for each respective battery. The four-item HAM-4-mTBI was calculated from the FGA and HiMAT using the individual item scores of Walk with Pivot Turn, Walk with Horizontal Head Turns, Fast Forward Walk, and Fast Backward Walk (12). Additionally, to capture a traditional measure of self-selected walking speed, straight-path gait speed was extracted from the ST and DT 1MW tests using validated and automated Mobility Lab software (APDM, a Clario Company, Portland, OR, USA).

#### Ecologically-Relevant Functional Tasks

Performance on the CAT was quantified as the total time to complete the course. Performance on the SUP was quantified using a throughput score (total accuracy score/total time), where points were awarded based on the Comstock method that allows for unlimited rounds, but heavily penalizes shooting the wrong target (i.e., shooting a friendly target is twice as bad as missing a hostile target). Each trial therefore had a total possible points [possible points = 2 * (Number of friendly targets) + 1 * (Number of hostile targets)] and an accuracy score [accuracy score = Possible points – 2 * (Number of tagged friendly targets) – 1 * (Number of untagged hostile targets)].(51) The final measure for performance on the SUP was the total throughput, defined as the sum of the total points divided by the sum of trial times 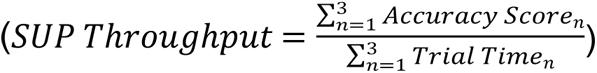.

### Statistical Analysis

To determine the diagnostic accuracy of turning outcomes to discriminate between healthy controls and people with mTBI, we first conducted a variable reduction procedure on all 152 primary and exploratory variables using lasso regression with 10-fold cross-validation to minimize the model deviance. Next, we constructed separate logistic regression models for each variable retained by the lasso regression to investigate the AUC for each turning variable individually. Then, we constructed ROC curves for a) the lasso model including all retained variables and b) the univariate logistic regression model considering each retained variable separately. For each ROC curve, we calculated the AUC and the 95% confidence interval (CI) of the AUC using bootstrapping with 10,000 iterations.

Since it is possible that retained variables all originate from separate tests (i.e., 1MW vs. mIAT vs. CTC), we further investigated the diagnostic accuracy of *individual tests* by running three separate lasso regressions, each with the same 10-fold cross-validation to minimize model deviance. Each lasso regression model included only variables from a single test (e.g., 1MW vs. mIAT vs. CTC), but included both ST and DT conditions. AUCs and 95% CIs were generated for each test using the same process described above.

To determine if turning outcomes have added value over standard clinical assessment batteries, forward stepwise logistic regression models were implemented using each clinical assessment as the base predictor. Variables were added to the base model in order of their univariate AUC (highest AUC added first). Separate models were fit for each clinical assessment (FGA, HiMAT, gait speed). Stopping criteria were determined using the Akaike Information Criteria (AIC). AUC values and 95% CIs were determined from the final model using bootstrapping with 10,000 iterations.

To assess the capacity of objective turning measures to predict performance in the ecologically-relevant functional tasks (CAT or SUP), partial correlation coefficients assessed the linear relationship between each variable and the performance outcome for the CAT (time to completion) and SUP (throughput score) while adjusting for age, body mass index (BMI), sex, mTBI status, and site. A 0.05 significance level with Benjamini-Hochberg false discovery rate (FDR) correction (52) was used throughout.

## RESULTS

Descriptive statistics for each variable and tests for between-group differences are presented in the supplemental material (Supplement B).

### Diagnostic Accuracy

A total of 22 of the possible 152 turning variables were retained following the lasso regression (Figure 2). The multivariate lasso model yielded an AUC (95% CI) of 0.92 (0.85, 0.96). Of the 22 retained turning variables, the average lap time during the ST CTC had the single largest AUC (95% CI) of 0.70 (0.58, 0.78). When lasso models were run as individual tests, the model initially including all 1MW outcomes yielded an AUC (95% CI) of 0.71 (0.61, 0.81) with 6 variables retained in the final model, the mIAT yielded an AUC (95% CI) of 0.78 (0.69, 0.86) with 8 retained variables, and the CTC yielded an AUC (95% CI) of 0.90 (0.84, 0.95) with 16 retained variables (Figure 3).

**Fig 2.**
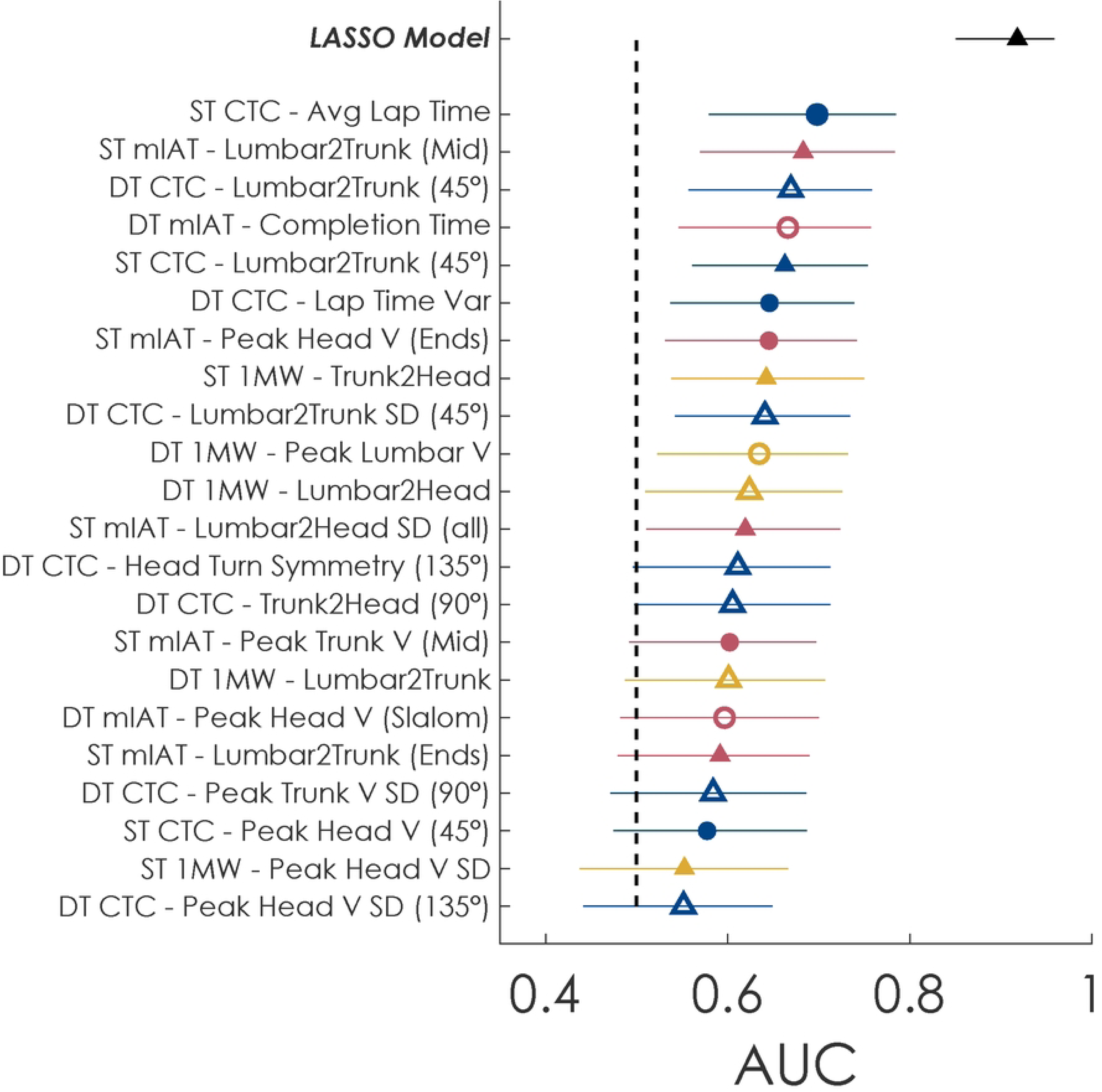
Areas under the reciever-operator characteristic curve (AUC) for the final lasso model (black) and univariate AUCs for each term included in the lasso model. Univariate AUCs are presented in different symbols for the type of variable (circle = speed, triangle = segmental coordination), different colors for the different tests (blue = complex turning course, yellow = modified Illinois Agility Test, red = one-minute walk test), and different fills for single- or dual-task (filled shape = single-task, empty shape = dual-task). Whiskers indicate the 95% confidence interval for each AUC.

**Fig 3.**
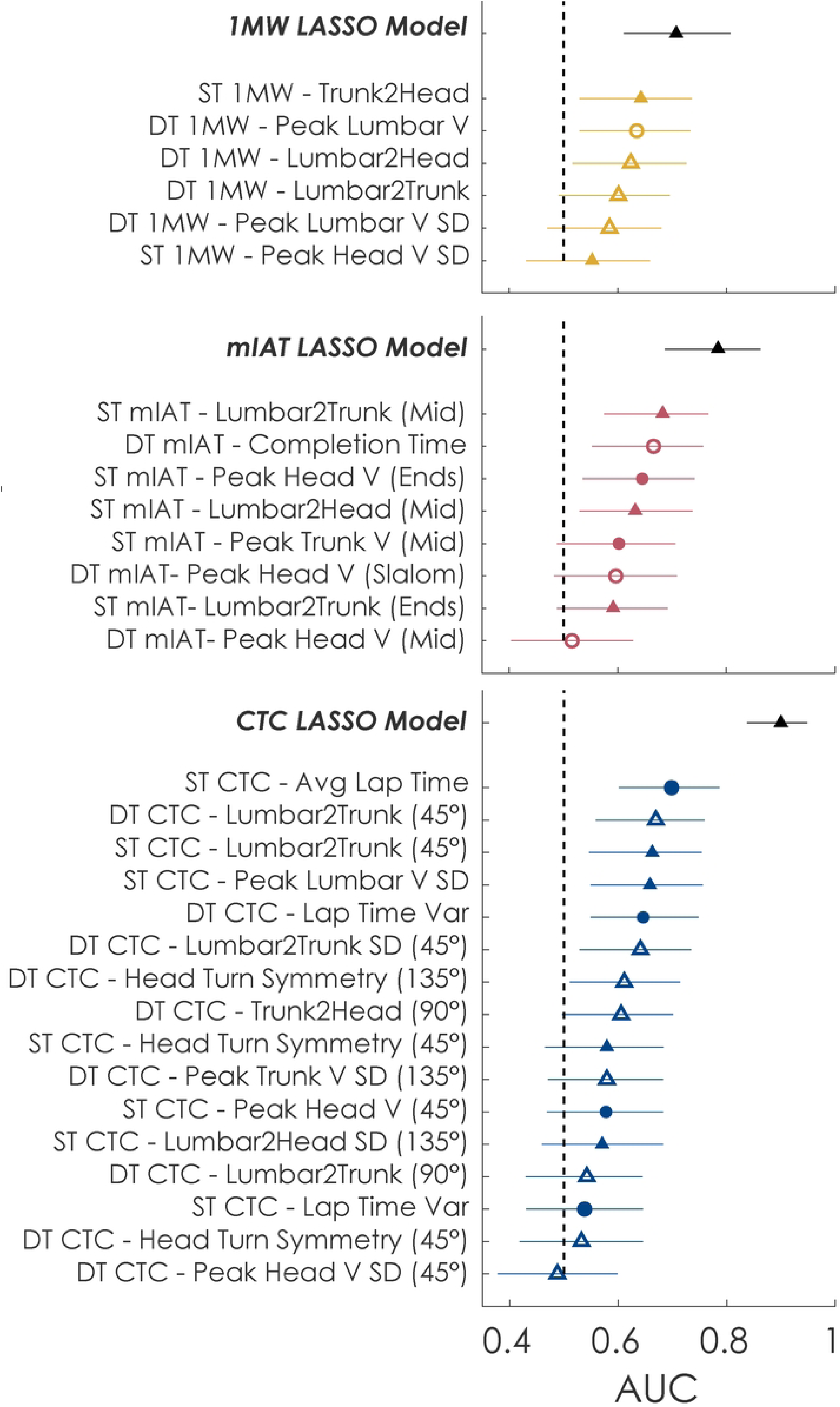
Areas under the reciever-operator characteristic curve (AUC) for the lasso models for individual tests. Each panel includes the final lasso model (black) and univariate AUCs for each term included in the lasso models. Univariate AUCs are presented in different symbols for the type of variable (circle = speed, triangle = segmental coordination), different colors for the different tests (blue = complex turning course, yellow = modified Illinois Agility Test, red = one-minute walk test), and different fills for single- or dual-task (filled shape = single-task, empty shape = dual-task). Whiskers indicate the 95% confidence interval for each AUC.

### Added Value of Clinic-Based Turning Tasks

The forward stepwise logistic models achieved stopping criteria (local minimum in AIC) after three turning variables for the FGA, HiMAT, and HAM-4-mTBI models (Table 2). The model based on the FGA achieved a final AUC (95% CI) of 0.80 (0.70, 0.87). The model based on the HiMAT achieved a final AUC (95% CI) of 0.79 (0.70, 0.86). The model based on the HAM-4-mTBI achieved a final AUC (95% CI) of 0.81 (0.73, 0.88). Stopping criteria was achieved after the addition of four turning variables for ST and DT gait speed, with final AUCs (95% CI) of 0.82 (0.72, 0.88) and 0.82 (0.73, 0.89), respectively. Comparatively, the AUC (95% CI) was 0.68 (0.57, 0.77) for the FGA, 0.65 (0.53, 0.74) for the HiMAT, 0.71 (0.61, 0.80) for the HAM-4-mTBI, 0.63 (0.52, 0.74) for ST gait speed, and 0.64 (0.53, 0.75) for DT gait speed.

**Table 2.** Results of the area under the receiver operator characteristic curve (AUC) from the stepwise logistic regression models to evaluate the added value of turning variables over standard mobility assessments. Base models included the Functional Gait Assessment (FGA), High-level Mobility Assessment Tool (HiMAT), 4-Item Hybrid Assessment of Mobility for mild Traumatic Brain Injury (HAM-4-mTBI), single-task (ST) gait speed, and dual-task (DT) gait speed. Each model included the following terms added, in order, to the base model: average lap time for the ST complex turning course (CTC); lumbar-to-trunk coordination during the middle turn on the ST modified Illinois Agility Test (mIAT); lumbar-to-trunk coordination during 45° turns of the DT CTC; and completion time for the DT mIAT.

| Model | FGA |  |  | HiMAT |  |  | HAM-4-mTBI |  |  | ST Gait Speed |  |  | DT Gait Speed |  |  |
| --- | --- | --- | --- | --- | --- | --- | --- | --- | --- | --- | --- | --- | --- | --- | --- |
|  | AUC | 95% CI |  | AUC | 95% CI |  | AUC | 95% CI |  | AUC | 95% CI |  | AUC | 95% CI |  |
| Base | 0.68 | 0.57 | 0.77 | 0.65 | 0.53 | 0.74 | 0.71 | 0.61 | 0.80 | 0.63 | 0.52 | 0.74 | 0.64 | 0.53 | 0.75 |
| Base+1 | 0.75 | 0.64 | 0.84 | 0.74 | 0.63 | 0.83 | 0.78 | 0.68 | 0.86 | 0.70 | 0.59 | 0.79 | 0.70 | 0.60 | 0.79 |
| Base+2 | 0.78 | 0.67 | 0.86 | 0.79 | 0.70 | 0.86 | 0.80 | 0.70 | 0.87 | 0.73 | 0.63 | 0.82 | 0.73 | 0.63 | 0.82 |
| Base+3 | 0.80 | 0.70 | 0.87 | 0.81 | 0.72 | 0.88 | 0.81 | 0.73 | 0.88 | 0.77 | 0.67 | 0.84 | 0.77 | 0.66 | 0.84 |
| Base+4 | - | - | - | - | - | - | - | - | - | 0.82 | 0.72 | 0.88 | 0.82 | 0.73 | 0.89 |

### Association with Ecologically-Relevant Functional Tasks (CAT and SUP)

Descriptive statistics for the CAT and SUP are presented in the supplemental material (Supplement C). Better performance on the CAT was significantly associated with faster overall lap times of the ST and DT CTC (|r| = 0.53, p < 0.0001 and |r| = 0.47, p < 0.0001, respectively) and faster peak turning speed of the sternum and lumbar spine during the 1MW and CTC tasks (|r| = 0.27-0.42, p < 0.05) (Figure 4 and Table 3). Instrumented measures from ST and DT straight gait were similarly associated with CAT completion time (|r| = 0.30-0.51, p < 0.05), with DT gait speed having the strongest association. Performance on the CAT was not significantly associated with performance on the FGA, HiMAT, or HAM-4 (|r| = 0.19-0.24, p > 0.05).

**Figure 4.**
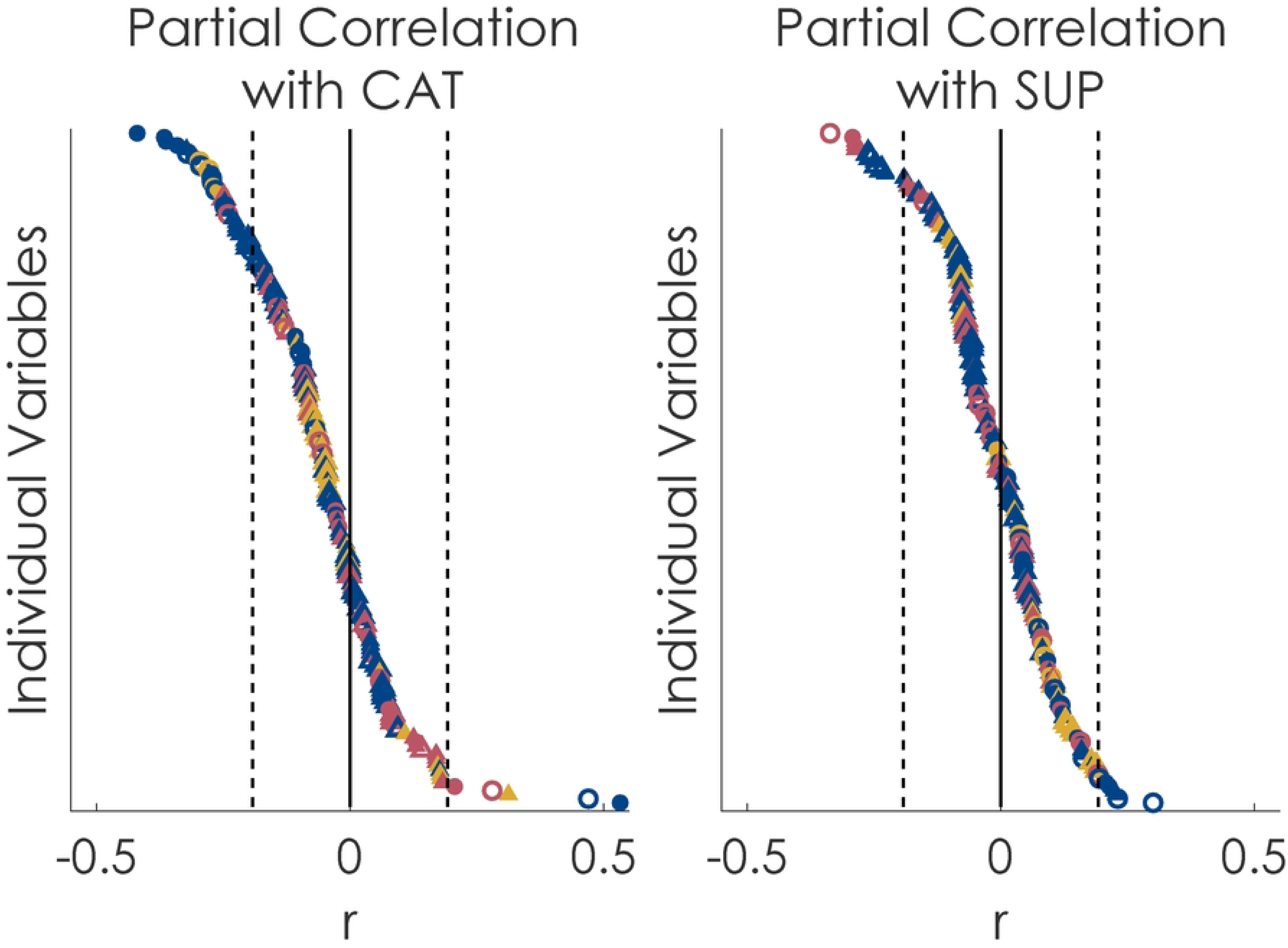
Partial Pearson correlation correficients for each turning variable obtained from the complex turning course (CTC), modified Illinois Agility Test (mIAT), and one-minute walk test (1MW) with completion time on the community ambulatory task (CAT; left) and simulated urban patrol (SUP; right). Correlation coefficients are shown in different symbols for the type of variable (circle = speed, triangle = segmental coordination), different colors for the different tests (blue = complex turning course, yellow = modified Illinois Agility Test, red = one-minute walk test), and different fills for single- or dual-task (filled shape = single-task, empty shape = dual-task).

**Table 3.** Partial correlation coefficients between variables from the clinic-based turning tasks, general measures of mobility, and the community ambulation task (CAT). All variables are ranked by magnitude of correlation coefficient. Variables from clinic-based turning tasks are noted with an * and only variables with significant associations (adjusted *p* < 0.05) are shown.

| | Partial Correlation Coefficient ($r$) | Adjusted $p$ value |
| --- | --- | --- |
| <u>Clinic-Based Turning Tasks</u> |  |  |
| ST CTC - Avg Lap Time* | 0.533 | <0.001 |
| DT CTC - Avg Lap Time* | 0.470 | <0.001 |
| ST CTC - Peak Lumbar V (90°)* | -0.420 | <0.001 |
| ST CTC - Peak Lumbar V (45°)* | -0.366 | 0.003 |
| ST CTC - Peak Lumbar V (135°)* | -0.363 | 0.003 |
| ST CTC - Peak Trunk V (135°)* | -0.341 | 0.006 |
| ST CTC - Peak Lumbar V SD (45°)* | -0.322 | 0.011 |
| DT CTC - Peak Lumbar V (90°)* | -0.321 | 0.011 |
| ST 1MW - Trunk2Head* | 0.312 | 0.013 |
| ST CTC - Peak Trunk V (90°)* | -0.308 | 0.015 |
| DT 1MW - Peak Lumbar V* | -0.296 | 0.021 |
| ST CTC - Peak Trunk V (135°)* | -0.295 | 0.021 |
| DT mIAT - Completion Time* | 0.280 | 0.033 |
| DT 1MW - Peak Trunk V* | -0.278 | 0.033 |
| ST CTC - Peak Trunk V (45°)* | -0.273 | 0.037 |
| DT CTC - Peak Trunk V (45°)* | -0.273 | 0.037 |
| DT CTC - Peak Lumbar V (135°)* | -0.272 | 0.037 |
| ST 1MW - Peak Lumbar V* | -0.270 | 0.038 |
| DT CTC - Peak Lumbar V (45°)* | -0.265 | 0.043 |
| <u>General Measures of Mobility</u> |  |  |
| Dual-task Gait Speed | -0.508 | <0.001 |
| Single-task Gait Speed | -0.455 | <0.001 |
| FGA Total Score | -0.235 | 0.083 |
| HiMAT Total Score | -0.228 | 0.095 |
| HAM-4-mTBI | -0.188 | 0.210 |

1MW = one-minute walk test; CTC = complex turning course; FGA = Functional Gait Assessment; HAM-4-mTBI = 4-Item Hybrid Assessment of Mobility for mild Traumatic Brain Injury; HiMAT = High-level Mobility Assessment Tool; mIAT = modified Illinois Agility Test; ST = single-task; DT = dual-task; SD = standard deviation

Better performance on the SUP was only significantly associated with faster DT mIAT times (|r| = 0.34, p = 0.031). The association between SUP performance and head-body coordination variability and peak trunk turning speed during the CTC approached but failed to reach significance (|r| = 0.29-0.30, p > 0.05; Figure 4 and Table 4). No clinical measure (FGA, HiMAT, HAM-4-mTBI) was significantly associated with SUP performance (|r| = 0.15-0.29, p > 0.05). Amongst clinical measures, the HiMAT score was most strongly associated with SUP performance (|r| = 0.29, p = 0.061).

**Table 4.** Partial correlation coefficients between variables from the clinic-based turning tasks (*), general measures of mobility, and the simulated urban patrol (SUP) task. All measures are ranked by magnitude of correlation coefficient. Instrumented turning measures are noted with an * and only instrumented turning measures with significant associations (adjusted *p* < 0.05) are shown.

| | Partial Correlation<br>Coefficient ($r$) | Adjusted $p$ value |
| --- | --- | --- |
| <u>Clinic-Based Turning Tasks</u> |  |  |
| Dual-task mIAT - Completion Time* | -0.336 | 0.031 |
| <u>General Measures of Mobility</u> |  |  |
| HiMAT Total Score | 0.291 | 0.061 |
| Dual-task Gait Speed | 0.243 | 0.136 |
| Single-task Gait Speed | 0.234 | 0.156 |
| HAM-4-mTBI | 0.166 | 0.432 |

*HAM-4-mTBI = 4-Item Hybrid Assessment of Mobility for mild Traumatic Brain Injury; HiMAT = High-level Mobility Assessment Tool; mIAT = modified Illinois Agility Test*

## DISCUSSION

This study sought to assess the clinical utility, including the diagnostic accuracy and associations with real-world performance, of objective turning measures from clinic-based turning tasks. Our results indicate that individual, objective turning measures from clinic-based turning tasks offer similar diagnostic capacities to standard clinical batteries such as the FGA, HiMAT, and HAM-4-mTBI. The value of instrumented clinic-based turning tasks, however, stems from the ability to capture multi-dimensional variables at once. Regression models using multiple variables from the same test yielded higher AUCs compared to standard clinical assessments (e.g., CTC AUC of 0.90 vs. FGA AUC of 0.68). Further, objective outcomes, including lap time from the ST CTC and segmental coordination from the ST mIAT and DT CTC provided added value by improving the AUC of the FGA, HiMAT, HAM-4-mTBI, and standard assessments of ST or DT gait speed. This superior diagnostic capacity and added value of clinic-based turning tasks, with time commitments (<10 minutes) that are similar or shorter than standard clinical batteries, suggests instrumented, clinic-based turning tasks could improve clinical decisions, including RTD decisions. The military’s Progressive Return to Activity: Primary Care for Acute Concussion Management outlines a physical and cognitive RTD screening “to objectively measure readiness for return to duty.” However, the physical RTD screening only requires two minutes of supervised aerobic activity at an exertion rate of 16 or greater on the Borg Rate of Perceived Exertion scale. Overt symptom provocation is the only metric for determining readiness to progress to the cognitive RTD screening, the final step before returning a service member to full duty. Clinic-based turning tasks, especially when instrumented, could better inform readiness for RTD than a service member’s display of overt symptom provocation or self-report of worsening symptoms with exertion.

Objective turning variables were strongly associated with real-world function on the CAT or SUP, indicating a highly desirable feature for informing RTD decisions. A summary of key stakeholders, including rehabilitation specialists and military command leaders, reported that RTD decisions are primarily based on whether an individual can complete the required duties of their position.(53) Yet, few clinical assessments accurately reflect the demands of daily living, especially specialized military tasks.(53) Out of the standard clinical assessments, only ST and DT gait speed associated with the CAT, while 19 turning measures were associated with CAT performance. Given the nature of the CAT (walking and navigating through a building), it is unsurprising that gait speed was associated with CAT performance. However, lap time from the ST CTC exhibited a slightly stronger association with CAT performance. This result indicates that assessments of ambulatory turning may be more relevant to a person’s daily life than straight-line walking. Similarly, completion time on the DT mIAT was the only variable associated with performance on the SUP; no standard clinical assessments were associated with this military-relevant activity. These results suggest the maximum running and turning speed during the mIAT better reflects high-demand simulated battle drills and may be more important than standard clinical assessments for extrapolating an individual’s performance in combat. Movements during the mIAT may be similar to individual movement techniques service members must perform, like the 3-5 second rush (moving under fire).

Using inertial sensors to capture multiple objective measures of turning enabled individual tasks to achieve high diagnostic capacity, but isolated variables of CTC lap time and mIAT completion time had the strongest clinical utility. Average lap time from the ST CTC had the largest AUC of any clinic-based turning variable and the strongest association with CAT performance. Similarly, DT mIAT completion time had the fourth highest AUC and was the only variable associated with SUP performance. Both these measures reflect an overall performance on the CTC and mIAT task, respectively. While other variables such as peak turning speed and segmental coordination are isolated to specific turns within a task, these measures of lap / completion time include all aspects of the task. Specific features, such as the anticipatory adjustments when initiating or terminating a turn, and the walking or running in between turns, were not quantified by our selected variable set and may be important to understanding an individual’s deficits, particularly during daily life. Notably, these lap time and completion time variables are reliable and do not require instrumentation (42). Both CTC lap time and mIAT completion time could be obtained from a stopwatch for rapid implementation in military and civilian clinics without access to inertial sensors.

A final goal of this study was to provide recommendations for future clinical adoption. Based on the available evidence from this study and prior studies on test-retest reliability, the CTC likely offers the best clinical utility. The CTC yielded the largest AUC values as a combined test, the individual variables with the largest univariate AUCs (ST CTC average lap time), and the individual variables with the strongest association with daily living. Combined, this suggests that the CTC may be a valuable addition to clinical mTBI evaluations, particularly if objective measures from inertial sensors are available to generate the full variable set. However, before CTC variables can be used for clinical decisions, future research should examine changes to the CTC over the course of rehabilitation and whether better performance on the CTC associates with RTD outcomes such as faster RTD in the military population, return to sport in athletic populations, risk for musculoskeletal injuries that are common after mTBI (54–59), and overall performance in one’s military occupational specialty.

While the CTC may offer the best clinical utility out of the tasks examined here, it is unlikely to be a panacea for RTD assessments. Complex and ecologically valid assessment techniques incorporating DT and multitask methods may prove useful in validating return-to-activity requirements in civilian and military populations as they more closely mimic real-world activity and are a step beyond single domain measures of impairment that do not capture the full picture of function (14). There are trade-offs to using more complex functional performance testing for return to activity decisions based on environment (e.g., deployed or garrison environment for military) and timeframe or operational needs for decisions to be made. Recently developed assessments like the Portable Warrior Test of Tactical Agility (POWAR-TOTAL) may be useful for testing military populations with combat roles, given its ability to discriminate service members with mTBI from controls and its responsiveness to rehabilitation (16, 60). However, such tasks may not be relevant for individuals with non-combat roles, including civilian populations. While the POWAR-TOTAL established construct validity using the HiMAT (60), the present CAT results suggest that the HiMAT lacks construct validity for functional tasks for civilians. Rather than reliance on a single assessment, the continued development of multiple complementary, short, and clinically useful assessments that serve as a ‘menu’ based on the patient’s needs may be ideal.

### Limitations

A primary limitation of this study is the inclusion of only civilians with symptoms persisting beyond 3 weeks post-injury; military service members were not included. This study’s patient population was selected because they represent individuals who seek rehabilitation and where return to work or sport decisions can be most complicated and were accessible to the study team at each site. However, these patients may not represent all individuals, including active-duty military personnel, law enforcement officers, or other tactical athletes for whom the SUP task is most ecologically-relevant. This limitation may explain the relatively weak associations we observed between the SUP and all variables, especially when compared to the CAT. A second limitation of this work was not considering mechanisms of injury in our analyses. It is possible that differences in the mechanisms of injury (e.g., motor-vehicle accident vs. sport-related vs. fall, etc.) may influence performance differently. Finally, this study did not assess other aspects that may influence an individual’s ability to return to pre-injury activity, such as their psychological readiness. Prior work in athletes has identified transient changes in psychological readiness, including a lack of confidence in their ability to handle the demands of competition (61, 62). While objective measures of turning offer a valuable clinical tool for measuring behavioral outcomes, RTD decisions should also evaluate the individual’s confidence to handle the task demands, which may also affect their readiness to return.

## CONCLUSION

Simple turning outcomes can discriminate individuals with persistent mTBI symptoms from healthy controls as well as standard clinical batteries. Instrumented outcomes that quantify intersegmental coordination during these turning tasks provide even more benefit and can increase the diagnostic accuracy of these tests. For the civilian participants in this study, turning outcomes were strongly associated with real-world ambulation and military-relevant battle drills; standard clinical assessments exhibited weaker, or non-significant, associations with these CAT and SUP tasks. Predicting real-world performance requires assessments that match the cognitive and physical demands of the operational environment. Future work should quantify the ambulatory demands of real-world tasks (such as ecological assessments during training tasks performed in the field) and compare them to normative performance on clinically-feasible turning tasks to further establish their validity.

## DATA AVAILABILITY

Data is available through the Federal Interagency Traumatic Brain Injury Research (FITBIR) Informatics System (doi.org/10.23718/study/390).

## ACKNOWLEDGEMENTS

The authors wish to acknowledge Ben Cassidy and Ryan Pelo, PT, DPT from the University of Utah, Lindsey Lee and Josh Koch from Oregon Health & Science University, Patrick Michielutti PT, DPT and Max Klaiman from Courage Kenny Research Center-Allina Health, as well as Holly Richard, PT, DPT and CPT Stefanie Faull, PT, DPT who collected data at Fort Sam Houston. Clinical Trials: https://clinicaltrials.gov/study/NCT03892291.

## DISCLOSURES / CONFLICTS OF INTEREST

The views expressed in this manuscript are those of the authors and do not necessarily represent the official policy or position of the U.S. Army Medical Center of Excellence, the U.S. Army Training and Doctrine Command, Department of the Army, Department of Defense, or any other U.S. Government agency. The authors declare that the research was conducted in the absence of any commercial or financial relationships that could be construed as a potential conflict of interest.

## FUNDING

This work was supported by the Assistant Secretary of Defense for Health Affairs endorsed by the Department of Defense, through the Congressionally Directed Medical Research Program under Award No. W81XWH1820049 (LK). An integrated SQL database at Oregon Health & Science University has housed all the data and is supported by the Oregon Clinical and Translational Research Institute funded by a grant from the National Center for Advancing Translational Sciences (NCATS), National Institutes of Health, through Grant Award Number UL1TR002369.

## Notes

### Competing Interest Statement

The authors have declared no competing interest.

### Clinical Trial

NCT03892291

### Clinical Protocols

https://clinicaltrials.gov/study/NCT03892291

### Author Declarations

The study was approved by the University of Utah (IRB_00114616), Oregona Health & Science University (STUDY00018749), and Allina Health (00002425).

## REFERENCES

1. Fino PC, Dibble LE, Wilde EA, Fino NF, Johnson P, Cortez MM, et al. Sensory Phenotypes for Balance Dysfunction After Mild Traumatic Brain Injury. Neurology. 2022;99(5):e521–35.

2. Haran FJ, Slaboda JC, King LA, Wright WG, Houlihan D, Norris JN. Sensitivity of the Balance Error Scoring System and the Sensory Organization Test in the Combat Environment. J Neurotrauma. 2016;33(7):705–11.

3. Buttner F, Howell DR, Ardern CL, Doherty C, Blake C, Ryan J, et al. Concussed athletes walk slower than non-concussed athletes during cognitive-motor dual-task assessments but not during single-task assessments 2 months after sports concussion: a systematic review and meta-analysis using individual participant data. Br J Sports Med. 2020;54(2):94–101.

4. Fino PC, Parrington L, Pitt W, Martini DN, Chesnutt JC, Chou LS, et al. Detecting gait abnormalities after concussion or mild traumatic brain injury: A systematic review of single-task, dual-task, and complex gait. Gait Posture. 2018;62:157–66.

5. Howell DR, Beasley M, Vopat L, Meehan WP, 3rd. The Effect of Prior Concussion History on Dual-Task Gait following a Concussion. J Neurotrauma. 2017;34(4):838–44.

6. Howell DR, Osternig LR, Chou LS. Single-task and dual-task tandem gait test performance after concussion. Journal of science and medicine in sport / Sports Medicine Australia. 2017;20(7):622–6.

7. Lee H, Sullivan SJ, Schneiders AG. The use of the dual-task paradigm in detecting gait performance deficits following a sports-related concussion: a systematic review and meta-analysis. Journal of science and medicine in sport / Sports Medicine Australia. 2013;16(1):2–7.

8. Register-Mihalik J, Littleton A, Guskiewicz K. Are Divided Attention Tasks Useful in the Assessment and Management of Sport-Related Concussion? Neuropsychol Rev. 2013;23(4):300–13.

9. Wrisley DM, Marchetti GF, Kuharsky DK, Whitney SL. Reliability, internal consistency, and validity of data obtained with the functional gait assessment. Phys Ther. 2004;84(10):906–18.

10. Williams G, Pallant J, Greenwood K. Further development of the High-level Mobility Assessment Tool (HiMAT). Brain Inj. 2010;24(7-8):1027–31.

11. Williams GP, Greenwood KM, Robertson VJ, Goldie PA, Morris ME. High-Level Mobility Assessment Tool (HiMAT): interrater reliability, retest reliability, and internal consistency. Phys Ther. 2006;86(3):395–400.

12. Fino PC, Michielutti PG, Pelo R, Parrington L, Dibble LE, Hoppes CW, et al. A Hybrid Assessment of Clinical Mobility Test Items for Evaluating Individuals With Mild Traumatic Brain Injury. Journal of neurologic physical therapy : JNPT. 2023;47(2):84–90.

13. Franchignoni F, Horak F, Godi M, Nardone A, Giordano A. Using psychometric techniques to improve the Balance Evaluation Systems Test: the mini-BESTest. Journal of rehabilitation medicine. 2010;42(4):323–31.

14. Scherer MR, Weightman MM, Radomski MV, Davidson LF, McCulloch KL. Returning service members to duty following mild traumatic brain injury: exploring the use of dual-task and multitask assessment methods. Phys Ther. 2013;93(9):1254–67.

15. Weightman MM, McCulloch KL, Radomski MV, Finkelstein M, Cecchini AS, Davidson LF, et al. Further Development of the Assessment of Military Multitasking Performance: Iterative Reliability Testing. PLoS One. 2017;12(1):e0169104.

16. McCulloch KL, Oh AS, Cecchini AS, Zhang W, Harrison C, Favorov O. Validity and Responsiveness of the Portable Warrior Test of Tactical Agility After Rehabilitation in Service Members With Mild Traumatic Brain Injury. Phys Ther. 2023;103(11).

17. Glaister BC, Bernatz GC, Klute GK, Orendurff MS. Video task analysis of turning during activities of daily living. Gait Posture. 2007;25(2):289–94.

18. Xu D, Carlton LG, Rosengren KS. Anticipatory postural adjustments for altering direction during walking. J Mot Behav. 2004;36(3):316–26.

19. Taylor M, Dabnichki P, Strike S. A three-dimensional biomechanical comparison between turning strategies during the stance phase of walking. Human Movement Science. 2005;24(4):558–73.

20. Imai T, Moore ST, Raphan T, Cohen B. Interaction of the body, head, and eyes during walking and turning. Exp Brain Res. 2001;136(1):1–18.

21. Raphan T, Imai T, Moore ST, Cohen B. Vestibular compensation and orientation during locomotion. Annals of the New York Academy of Sciences. 2001;942:128–38.

22. Bernardin D, Kadone H, Bennequin D, Sugar T, Zaoui M, Berthoz A. Gaze anticipation during human locomotion. Exp Brain Res. 2012;223(1):65–78.

23. Grasso R, Prevost P, Ivanenko YP, Berthoz A. Eye-head coordination for the steering of locomotion in humans: an anticipatory synergy. Neurosci Lett. 1998;253(2):115–8.

24. Hollands MA, Patla AE, Vickers JN. "Look where you’re going!": gaze behaviour associated with maintaining and changing the direction of locomotion. Exp Brain Res. 2002;143(2):221–30.

25. Stuart S, Parrington L, Morris R, Martini DN, Fino PC, King LA. Gait measurement in chronic mild traumatic brain injury: A model approach. Hum Mov Sci. 2020;69:102557.

26. Powell D, Godfrey A, Parrington L, Campbell KR, King LA, Stuart S. Free-living gait does not differentiate chronic mTBI patients compared to healthy controls. Journal of neuroengineering and rehabilitation. 2022;19(1):49.

27. Fino PC, Parrington L, Walls M, Sippel E, Hullar TE, Chesnutt JC, et al. Abnormal Turning and Its Association with Self-Reported Symptoms in Chronic Mild Traumatic Brain Injury. J Neurotrauma. 2018;35(10):1167–77.

28. Fino PC, Nussbaum MA, Brolinson PG. Locomotor deficits in recently concussed athletes and matched controls during single and dual-task turning gait: preliminary results. Journal of neuroengineering and rehabilitation. 2016;13(1):65.

29. Ellis MJ, Cordingley D, Vis S, Reimer K, Leiter J, Russell K. Vestibulo-ocular dysfunction in pediatric sports-related concussion. Journal of neurosurgery Pediatrics. 2015;16(3):248–55.

30. Kolev OI, Sergeeva M. Vestibular disorders following different types of head and neck trauma. Functional neurology. 2016;31(2):75–80.

31. Mucha A, Collins MW, Elbin RJ, Furman JM, Troutman-Enseki C, DeWolf RM, et al. A Brief Vestibular/Ocular Motor Screening (VOMS) assessment to evaluate concussions: preliminary findings. Am J Sports Med. 2014;42(10):2479–86.

32. Fino PC, Lockhart TE, Fino NF. Corner height influences center of mass kinematics and path trajectory during turning. J Biomech. 2015;48(1):104–12.

33. Hollands KL, Agnihotri D, Tyson SF. Effects of dual task on turning ability in stroke survivors and older adults. Gait Posture. 2014;40(4):564–9.

34. Fino PC, Wilhelm J, Parrington L, Stuart S, Chesnutt JC, King LA. Inertial Sensors Reveal Subtle Motor Deficits When Walking With Horizontal Head Turns After Concussion. The Journal of head trauma rehabilitation. 2019;34(2):E74–E81.

35. King LA, Horak FB, Mancini M, Pierce D, Priest KC, Chesnutt J, et al. Instrumenting the balance error scoring system for use with patients reporting persistent balance problems after mild traumatic brain injury. Arch Phys Med Rehabil. 2014;95(2):353–9.

36. King LA, Mancini M, Fino PC, Chesnutt J, Swanson CW, Markwardt S, et al. Sensor-Based Balance Measures Outperform Modified Balance Error Scoring System in Identifying Acute Concussion. Ann Biomed Eng. 2017.

37. Fino PC, Weightman MM, Dibble LE, Lester ME, Hoppes CW, Parrington L, et al. Objective Dual-Task Turning Measures for Return-to-Duty Assessment After Mild Traumatic Brain Injury: The ReTURN Study Protocol. Front Neurol. 2020;11:544812.

38. Buckley TA, Oldham JR, Caccese JB. Postural control deficits identify lingering post-concussion neurological deficits. J Sport Health Sci. 2016;5(1):61–9.

39. Riemann BL, Guskiewicz KM. Effects of mild head injury on postural stability as measured through clinical balance testing. J Athl Train. 2000;35(1):19–25.

40. Gera G, Chesnutt J, Mancini M, Horak FB, King LA. Inertial Sensor-Based Assessment of Central Sensory Integration for Balance After Mild Traumatic Brain Injury. Mil Med. 2018;183(suppl_1):327–32.

41. Berg K, Wood-Dauphinee S, Williams JI. The Balance Scale: reliability assessment with elderly residents and patients with an acute stroke. Scand J Rehabil Med. 1995;27(1):27–36.

42. Weston AR, Antonellis P, Fino PC, Hoppes CW, Lester ME, Weightman MM, et al. Quantifying Turning Tasks with Wearable Sensors: A Reliability Assessment. Phys Ther. 2023;104(2), pzad134.

43. VA/DoD Clinical Practice Guideline for the Management and Rehabiliation of Post-Acute Mild Traumatic Brain Injury. www.healthquality.va.gov/guidelines/Rehab/mtbi: Department of Veterans Affairs and Department of Defense; 2021 June 2021.

44. Patricios JS, Schneider KJ, Dvorak J, Ahmed OH, Blauwet C, Cantu RC, et al. Consensus statement on concussion in sport: the 6th International Conference on Concussion in Sport-Amsterdam, October 2022. Br J Sports Med. 2023;57(11):695–711.

45. Lieberman HR, Bathalon GP, Falco CM, Morgan CA, 3rd, Niro PJ, Tharion WJ. The fog of war: decrements in cognitive performance and mood associated with combat-like stress. Aviat Space Environ Med. 2005;76(7 Suppl):C7–14.

46. Palmer CJ, Bigelow C, Van Emmerik RE. Defining soldier equipment trade space: load effects on combat marksmanship and perception-action coupling. Ergonomics. 2013;56(11):1708–21.

47. Antonellis P, Weightman MM, Fino PC, Chen S, Lester ME, Hoppes CW, et al. Relation Between Cognitive Assessment and Clinical Physical Performance Measures After Mild Traumatic Brain Injury. Arch Phys Med Rehabil. 2023.

48. Loyd BJ, Dibble LE, Weightman MM, Pelo R, Hoppes CW, Lester M, et al. Volitional Head Movement Deficits and Alterations in Gait Speed Following Mild Traumatic Brain Injury. The Journal of head trauma rehabilitation. 2023;38(3):E223–E32.

49. Parrington L, King LA, Hoppes CW, Klaiman MJ, Michielutti P, Fino PC, et al. Exploring Vestibular Ocular Motor Screening in Adults With Persistent Complaints After Mild Traumatic Brain Injury. The Journal of head trauma rehabilitation. 2022;37(5):E346–E54.

50. Parrington L, King LA, Weightman MM, Hoppes CW, Lester ME, Dibble LE, et al. Between-site equivalence of turning speed assessments using inertial measurement units. Gait Posture. 2021;90:245–51.

51. USPSA Competition Rules, 9.2.2 (2019).

52. Benjamini Y, Hochberg Y. Controlling the False Discovery Rate - a Practical and Powerful Approach to Multiple Testing. J R Stat Soc B. 1995;57(1):289–300.

53. Radomski MV, Weightman MM, Davidson LF, Finkelstein M, Goldman S, McCulloch K, et al. Development of a measure to inform return-to-duty decision making after mild traumatic brain injury. Mil Med. 2013;178(3):246–53.

54. Brooks MA, Peterson K, Biese K, Sanfilippo J, Heiderscheit BC, Bell DR. Concussion Increases Odds of Sustaining a Lower Extremity Musculoskeletal Injury After Return to Play Among Collegiate Athletes. Am J Sports Med. 2016;44(3):742–7.

55. Lynall RC, Mauntel TC, Pohlig RT, Kerr ZY, Dompier TP, Hall EE, et al. Lower Extremity Musculoskeletal Injury Risk After Concussion Recovery in High School Athletes. J Athl Train. 2017;52(11):1028–34.

56. McPherson AL, Nagai T, Webster KE, Hewett TE. Musculoskeletal Injury Risk After Sport-Related Concussion: A Systematic Review and Meta-analysis. Am J Sports Med. 2019;47(7):1754–62.

57. Fino PC, Becker LN, Fino NF, Griesemer B, Goforth M, Brolinson PG. Effects of Recent Concussion and Injury History on Instantaneous Relative Risk of Lower Extremity Injury in Division I Collegiate Athletes. Clin J Sport Med. 2019;29(3):218–23.

58. Reneker JC, Babl R, Flowers MM. History of concussion and risk of subsequent injury in athletes and service members: A systematic review and meta-analysis. Musculoskelet Sci Pract. 2019;42:173–85.

59. Kardouni JR, Shing TL, McKinnon CJ, Scofield DE, Proctor SP. Risk for Lower Extremity Injury After Concussion: A Matched Cohort Study in Soldiers. J Orthop Sports Phys Ther. 2018;48(7):533–40.

60. Cecchini AS, Prim J, Zhang W, Harrison CH, McCulloch KL. The Portable Warrior Test of Tactical Agility: A Novel Functional Assessment That Discriminates Service Members Diagnosed With Concussion From Controls. Mil Med. 2023;188(3-4):e703–e10.

61. Crofts RM. Psychological Readiness to Return to Play After Concussion: The University of Utah; 2023.

62. van Ierssel J, Pennock KF, Sampson M, Zemek R, Caron JG. Which psychosocial factors are associated with return to sport following concussion? A systematic review. J Sport Health Sci. 2022;11(4):438–49.

